# Cross-platform metabolomics imputation using importance-weighted autoencoders

**DOI:** 10.1101/2025.03.06.25323475

**Authors:** Alexander Smith, Rui Pinto, Loukas Zagkos, Ioanna Tzoulaki, Paul Elliott, Abbas Dehghan

**Affiliations:** Department of Epidemiology and Biostatistics, School of Public Health, Imperial College London, London, UK; UK Dementia Research Institute at Imperial College London, Hammersmith Hospital, London, UK; MRC-NIHR BRC National Phenome Centre, Department of Digestion, Metabolism and Reproduction, Division of systems medicine, Imperial College London, London, UK; Medical Research Council Centre for Environment and Health, School of Public Health, Imperial College London, London, UK; Centre for Systems Biology, Biomedical Research Foundation of the Academy of Athens, Athens, Greece

## Abstract

**Background:** Metabolomics data are often generated through different analytical platforms and different methods of identification and quantification which makes their synthesis and large-scale replication challenging. To address this, we applied generative deep learning to impute metabolites assayed by Metabolon, a commonly used commercial platform, using metabolomic features acquired by an untargeted liquid chromatography-mass spectrometry (LC-MS) platform.

**Methods:** We utilised a subset of 979 samples from the Airwave Health Monitoring Study which were assayed by both Metabolon and National Phenome Centre at Imperial College (NPC) LC-MS assays to develop an ensemble of importance-weighted autoencoders (IWAEs) which can perform cross-platform metabolomics imputation between the two assays. Using the ensemble, we generated a Metabolon equivalent dataset in 2,971 additional Airwave samples that lacked prior Metabolon measurements. We conducted observational associations with two clinical outcomes, body mass index (BMI) and C-reactive protein (CRP). We validated the ensemble and imputed data by investigating the concordance of the observational associations. This was done using both the imputed Metabolon dataset and the measured metabolite levels by Metabolon, and NPC in the Airwave study and Nightingale platform in the UK Biobank.

**Results:** Our imputation ensemble generated samples highly correlated with their real values across all Metabolon metabolites within a held-out test set with a mean sample correlation of 0.61 (IQR 0.55-0.67). The well-imputed subset included 199 (22%) of the metabolites present in the real Metabolon dataset where the imputed values accounted for at least 55% of the original variance (R^2^ ≥ 0.55) and a minimal uncertainty (R^2^ variance ≤ 0.025). The subset included 43 metabolites not previously identified within our LC-MS platform. When comparing the associations of the real and imputed Metabolon metabolites with BMI and CRP, the standardised beta-coefficients were highly correlated (ρ = 0.93 for BMI and 0.89 for CRP) with minimal mean difference (0.005 (0.04) for BMI, 0.005 (0.04) for CRP). Similar concordance occurred between the imputed Metabolon metabolites and equivalent UK Biobank (mean difference -0.007 (0.05) for BMI, 0.01 (0.04) for CRP) and our LC-MS platform (mean difference -0.013 (0.04) for BMI, -0.019 (0.04) for CRP).

**Conclusion:** This methodological innovation offers a scalable and accurate method for cross-platform imputation which could allow for to aggregate individual-level metabolomics data from different epidemiological studies, replication findings or conduct meta-analyses.

## Introduction

Metabolomics, a critical field in systems biology, focuses on the comprehensive analysis of metabolites - important small molecules in biological systems. Metabolites can be exogenous, like nutrients and drugs, or endogenous, like cholesterol and triglycerides. They are vital building blocks for tissues and have roles as regulatory agents and messengers as well as many other functions. They help in energy production, protein synthesis, metabolism regulation, and other cellular functions, thereby maintaining an organism’s internal balance and health. The study of metabolic profiles provides insights into the metabolic pathways within an organism at a given time, enhancing our understanding of the biological interplay between physiological and pathological conditions. This understanding is crucial for advancing biomarker discovery, disease diagnosis, treatment monitoring and drug discovery^1–3^.

In recent years a growing number of large epidemiologic studies have acquired metabolomics data^4^ but often suffer from small sample sizes and/or limited metabolite coverage. Large studies may have ten to a hundred thousand samples but small metabolite coverage, e.g., UK Biobank lipoprotein/metabolite dataset^5^, while smaller ones contain hundreds to thousands of samples with large metabolite coverage, e.g. MESA^6^. The latter group of studies would greatly benefit from combining their data with additional cohorts to form a larger discovery panel or robustly externally validating their findings. However, the variability in metabolomics platforms, annotation and quantification prevents easily combining or comparing metabolomic features across studies. Several methods have been developed to combine LC-MS metabolomics datasets that utilize the same or similar platforms^7,8^ but to our knowledge, none have been published which entirely recreates one dataset from another.

An innovative solution is to treat the variables of each dataset as “missing” in the others. This solution is feasible given the high correlation between metabolites. Moreover, recent advances in applying deep latent variable models, such as importance-weighted autoencoders ^9^, provide a highly flexible approach to imputation in the complex, high dimensional datasets generated by metabolomics studies. The complex associations between metabolites caused by interconnected biological pathways can be captured by such deep learning models using their ability to model infinitely complex functions, including non-linear relationships. In addition, an importance-weighted autoencoder specifically developed for imputation has shown state-of-the-art imputation performance^10^ and, as far as we are aware, they have yet to be explored for cross-platform metabolomics imputation.

This study showcases the imputation of an entire dataset of the Metabolon platform from untargeted liquid chromatography – mass spectrometry (LC-MS) dataset acquired by the UK National Phenome Centre (NPC), using an ensemble importance-weighted autoencoder. We then validate the imputations by comparing the observational associations of the imputed metabolites in the imputed Metabolon dataset with body mass index (BMI) and C-Reactive protein (CRP), to the associations of the real Metabolon metabolites with the same outcomes. Such an innovative approach could be very beneficial, as many epidemiological studies have metabolomics datasets using different approaches and platforms. The suggested method allows for a one-to-one comparison of the metabolites for validation purposes or aggregate analyses across studies.

## Methods

### Data sources

#### Airwave

The Airwave Health Monitoring Study is a longitudinal cohort study on the UK police forces, with recruitment and baseline measurements done from 2005 to 2014, and includes medical, biomedical, occupational and lifestyle information. All participants were voluntary and provided with written informed consent. The Airwave Health Monitoring Study is approved by the National Health Service Multi-Site Research Ethics Committee (MREC/13/NW/0588). All methods followed the relevant guidelines and regulations.

Airwave samples were divided into two separate sample sets. The first set consisted of 3000 samples that utilised lithium heparin plasma samples and underwent UPLC-MS profiling analysis for lipids and small metabolites at the National Phenome Centre (NPC, Imperial College London, London, UK), hereafter known as the “NPC-Only dataset”. The second set consisted of 2250 ethylenediaminetetraacetic (EDTA) plasma samples and were analysed by Metabolon, Inc. (Morrisville, NC, USA) with 1000 of these samples also analysed by the NPC. Hereafter, the subset of 1000 individuals that included measurements by both platforms are known as the “Metabolon-NPC dataset”, whilst the remaining 1250 samples, are known as the “Metabolon-Only dataset”. NPC metabolomic features were matched using the “M2S” computational methodology published by our group^8^, yielding the same 3585 metabolomic features across the two NPC-based datasets, 914 of which were annotated (306 unique annotations). In both Metabolon datasets, 1148 metabolites were measured, comprising 828 annotated and 320 unknowns. Metabolon metabolites which were invariant across all samples or had more than 25% missing values been removed, leaving 915 metabolites. Data description, sample preparation, metabolite annotation and data processing have been described elsewhere for Airwave NPC^11^ and for Airwave Metabolon^12^. Outliers for all datasets were removed using local outlier factor ^13^ with default settings across all measured metabolites resulting in the removal of 67 samples across all datasets.

### Study overview

The overall analysis plan is shown in **Figure 1**. Using the Metabolon-NPC dataset, we identified five clusters of metabolites which grouped based on biological pathway / chemical class. Next, we split the Metabolon-NPC dataset samples into three subsets; training (n=706), validation (n=78) and held-out test set (n=195). The training and validation sets were used to train an importance-weighted autoencoder for each of the five metabolite clusters and an additional autoencoder with all metabolites and metabolomic features. These attempted to recreate the entire set of Metabolon measurements within the held-out test set samples based on greedy selection across all models, creating an imputed Metabolon dataset. To assess the quality of the imputations, we explored the R^2^ and R^2^ variance across imputation iterations within the test-set samples. The Metabolon metabolites imputed with a quality above a defined threshold in the held-out test set were selected, and their values were imputed in the NPC-Only dataset. Finally, we evaluated the quality of the generated NPC-Only imputed Metabolon dataset by comparing metabolite observational associations with body mass index and C-Reactive protein to similar associations with the real metabolite measurements by Metabolon, NPC and UK Biobank.

**Figure 1.**
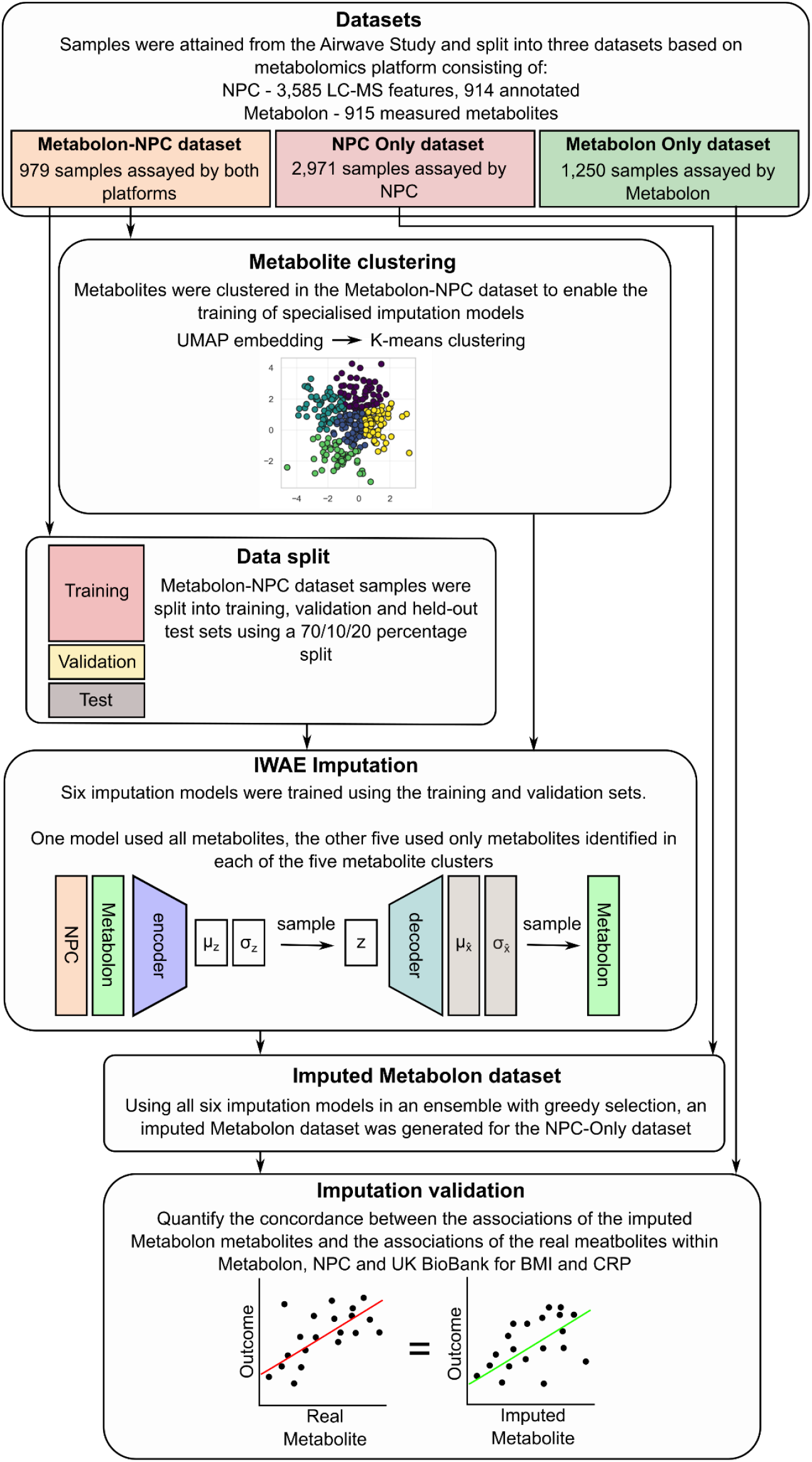
Schematic diagram of the study

**Figure 2.**
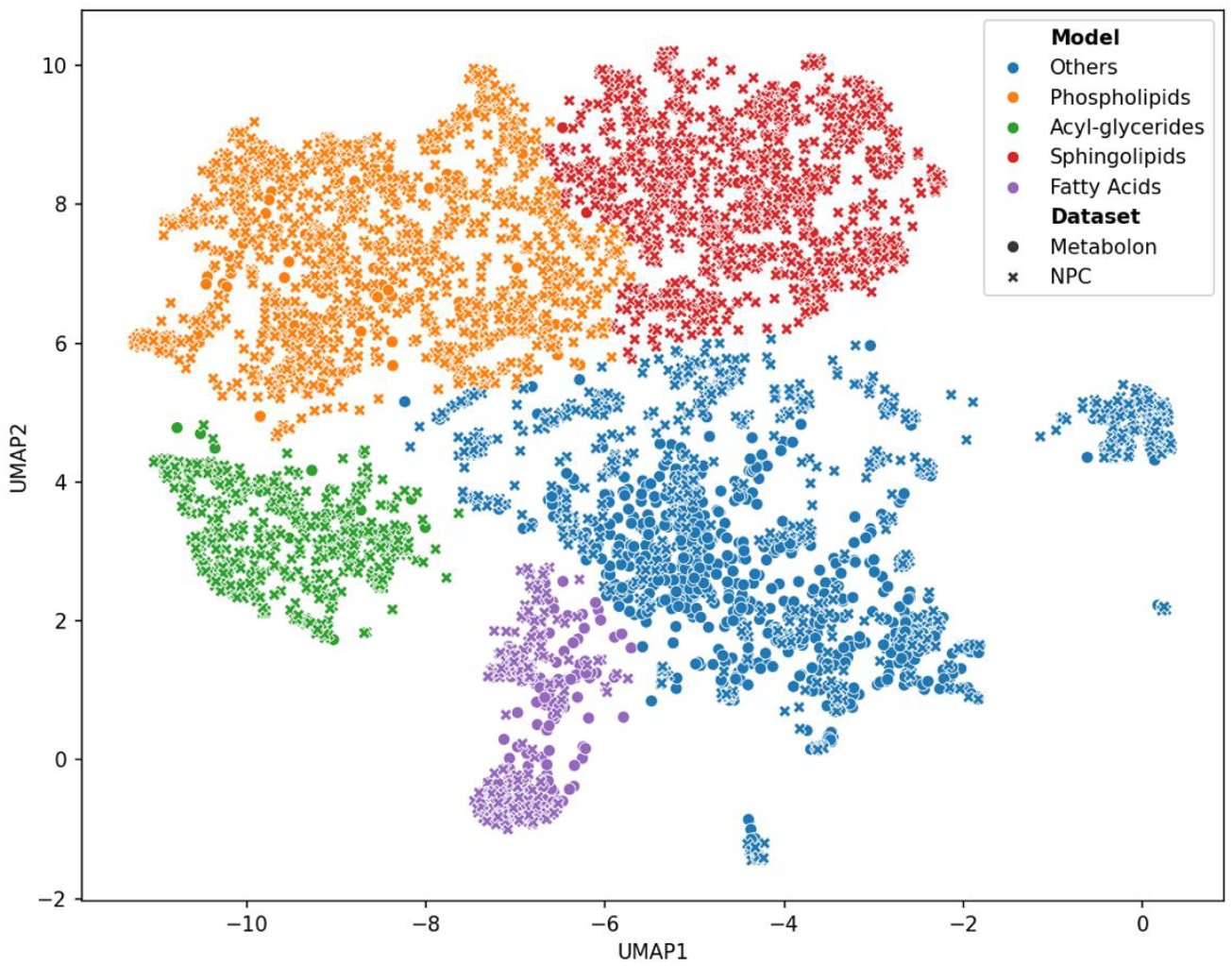
Metabolite clustering within the Metabolon-NPC dataset (N = 979) using both NPC and Metabolon metabolites

### Statistical analysis

#### Metabolite clustering

To identify clusters of metabolites within the Metabolon-NPC dataset, we used Uniform manifold approximation and projection (UMAP)^14^ for dimensionality reduction followed by a centroid-based clustering, a method that clusters items by similarity using central points for organization. UMAP was employed to map each of the NPC metabolomic features and Metabolon metabolites into the same 2-dimensional space, based on their values across the Metabolon-NPC dataset. UMAP was performed using cosine distance, two components, 35 neighbours and a minimum distance of 0.3. K-means clustering was used as the centroid-based clustering to identify the clusters of metabolites within the UMAP-embedded 2D space using the elbow method with silhouette score to determine the optimal number of clusters.

Clusters were labelled using their most predominant chemical compound class or biological pathway.

#### Metabolite imputation

The Metabolon-NPC dataset (N = 979) was separated into training, validation and held-out test set (70/10/20 split) prior to model training. To normalize the data, min-max scaling technique was applied, adjusting the value within the range of –1 to 1 using population estimates of mean and standard deviation calculated from only the training set. The core of the study involved training an importance-weighted autoencoder (IWAE) to impute the Metabolon metabolites by randomly masking single values and imputing a percentage of Metabolon metabolites during training.

Hyperparameters for each model were selected using coarse-to-fine grid searching including the type of IWAE^10,15^, the percentage of random masking, presence of mix-up regularisation^16^, dropout rate^17^, presence of batch normalisation^18^ and error function. Model performance was measured using the imputation error calculated after masking the entire set of Metabolon metabolites on the validation set with early stopping. After hyperparameter tuning, the final imputation model was trained using the combined training and validation set. This process was completed six times, once for each of the five metabolite clusters using only the metabolites and metabolomic features present in the cluster and once using all metabolites and metabolomic features.

For the evaluation phase, the model’s performance was tested by masking and imputing all Metabolon metabolites within the held-out test set. A selection process (greedy selection) determined the most appropriate imputation model for each metabolite based on the imputation performance within the validation set during model tuning. To assess the reliability and accuracy of the imputation, the test set underwent a total of fifty-one imputation cycles. The first cycle provided a baseline R^2^ performance for each metabolite, while the remaining fifty cycles were averaged to explore prediction uncertainty via R^2^ variance of the predicted vs observed values. Spearman’s rank correlation was used to compare the imputed values in the test set against the corresponding actual values. Finally, leveraging the finalised imputation model, an imputed Metabolon dataset was generated using the NPC-Only dataset (N = 2,971), employing the same greedy selection rules as applied to the test set.

#### Observational analysis

To further evaluate the accuracy of the imputations, metabolite associations with BMI and C-reactive protein (CRP) were examined using linear regression adjusted for age and sex.

Before fitting the models, both exposures and outcomes were standardised (mean = 0, SD = 1) before fitting the models. Only Metabolon metabolites which were imputed with an overall R^2^ ≥ 0.55 in the test set were tested for association using the imputed Metabolon (NPC-Only) dataset. This was selected as a reasonable trade-off point between metabolite imputation performance and uncertainty based on the test-set imputation performance (**Figure 3a**). The same set of metabolites was tested for association using the real Metabolon metabolites within the combined Metabolon-NPC and Metabolon-Only datasets (N = 2,229). Additionally, metabolites tested for association in the imputed Metabolon (NPC-Only) dataset which had a matching LC-MS annotation in the combined Metabolon-NPC and NPC-Only dataset (N = 3,950), or UK Biobank NMR (N = 110,068) were tested for associations (**Figure 6**).

**Figure 3.**
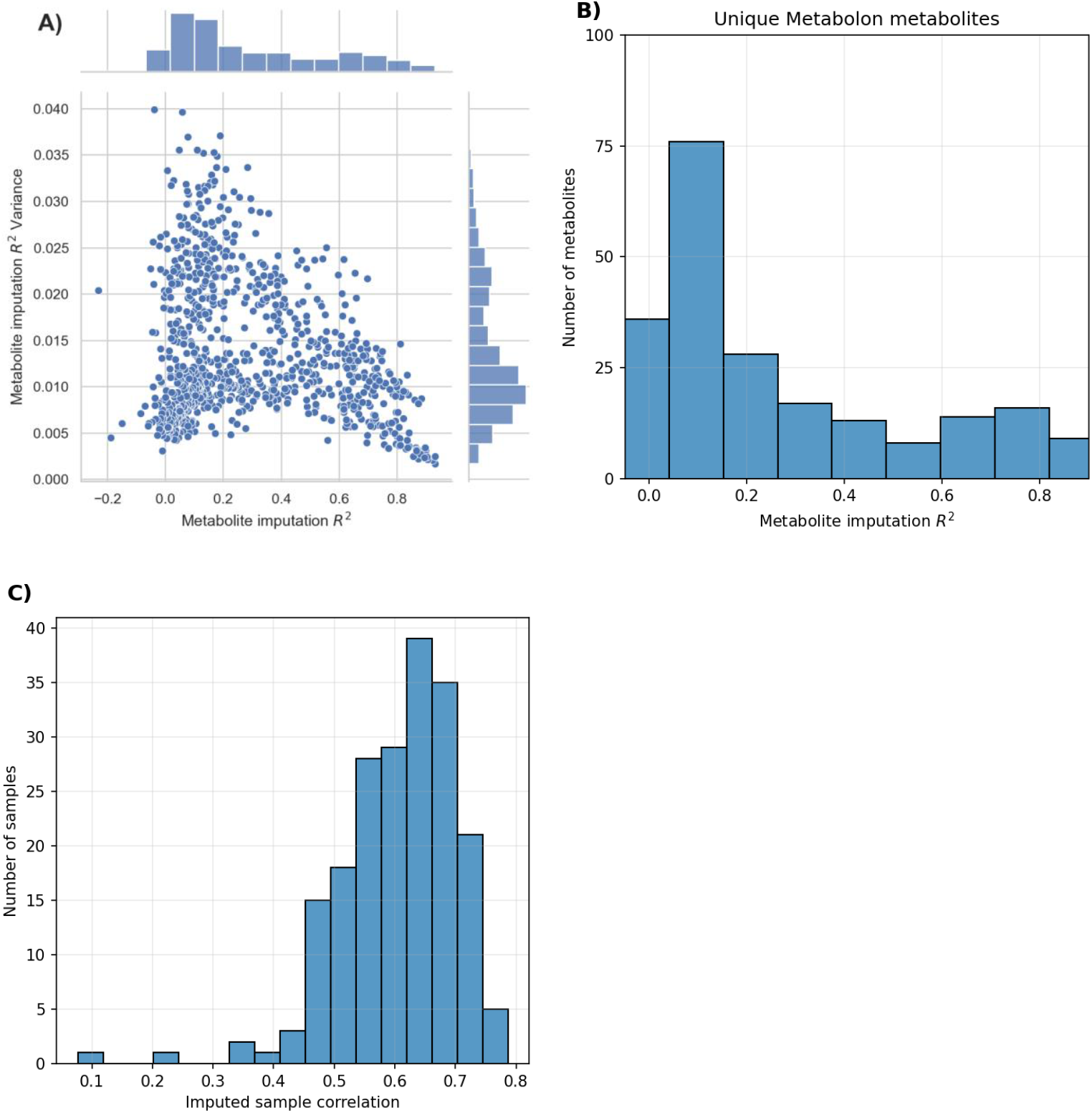
Imputation performance on the held-out test set; a) metabolite R^2^ vs R^2^ variance b) performance within the unique Metabolon metabolites c) imputed held-out test set sample correlation with real values

#### Software

NPC metabolomic features were matched across Airwave subsets using the “M2S” MATLAB (R2019b) package available at https://github.com/rjdossan/M2S^8^. Imputation was performed using TensorFlow v2.4.1^19^ and Python v3.7.9. Observational analyses and data processing were performed using sci-kit learn v0.23.2^20^.

## Results

Clustering the metabolites within the Metabolon-NPC dataset using the UMAP-embedded space identified five clusters. Based on their majority compound classes, four clusters were labelled Fatty acids, Sphingolipids, Phospholipids, and Acyl-glycerides. The fifth cluster was labelled Others for its lack of a predominant class (see **Supplementary Table 1** for full details). The clusters varied in size and composition ranging from a total of 356 metabolites for Fatty acids, including 134 Metabolon metabolites and 222 NPC metabolomic features, up to 503 metabolites for Acyl-glycerides (24 Metabolon).

IWAE imputation models, trained on each metabolite cluster, achieved low mean absolute error (MAE) on the test set samples (Fatty acids = 0.141, Sphingolipids = 0.140, Phospholipids = 0.132, Acyl-glycerides = 0.125 and Others = 0.190). The imputation model trained using the entire set of metabolites achieved a root mean squared error (RMSE) of 0.251 on the test set. The error rate (MAE or RMSE) for imputation on the combined training-validation set achieved results similar to the test set.

After greedy selection in the test set, the imputation of 644 metabolites achieved an R^2^ ≥ 0.1 (**Figure 3a**), of which 199 were equal to or surpassed an R^2^ of 0.55. Out of these 199 metabolites, 43 were unique to Metabolon compared to our LC-MS platform (**Figure 3b**). Uncertainty of imputation showed an inverse linear relationship with overall imputation performance (**Figure 3a**). The mean R^2^ variance for metabolites imputed with overall R^2^ ≥ 0.1 was 0.014, which drops to 0.009 for metabolites with overall R^2^ ≥ 0.55.

Metabolon categorises metabolites into superclasses based on their compound class. Imputation performance varied greatly across superclasses (**Figure 4**). The “Lipid” superclass was the best imputed with a mean R^2^ = 0.49, although it had the largest variance in performance (SD = 0.25). Xenobiotics (mean R^2^ = 0.13, SD = 0.15), Amino acids, (0.14, 0.14) Peptides (0.22, 0.15) and Cofactors and Vitamins (0.24, 0.25) had a combined total of 35 metabolites which imputed over an R^2^ of 0.3 but overall, they had low imputation performance. Carbohydrates, Energy and Nucleotide metabolites had no outstanding metabolites with poor mean R^2^ = 0.11 (SD = 0.08), 0.16 (SD = 0.08) and 0.11 (SD = 0.10) respectively.

**Figure 4.**
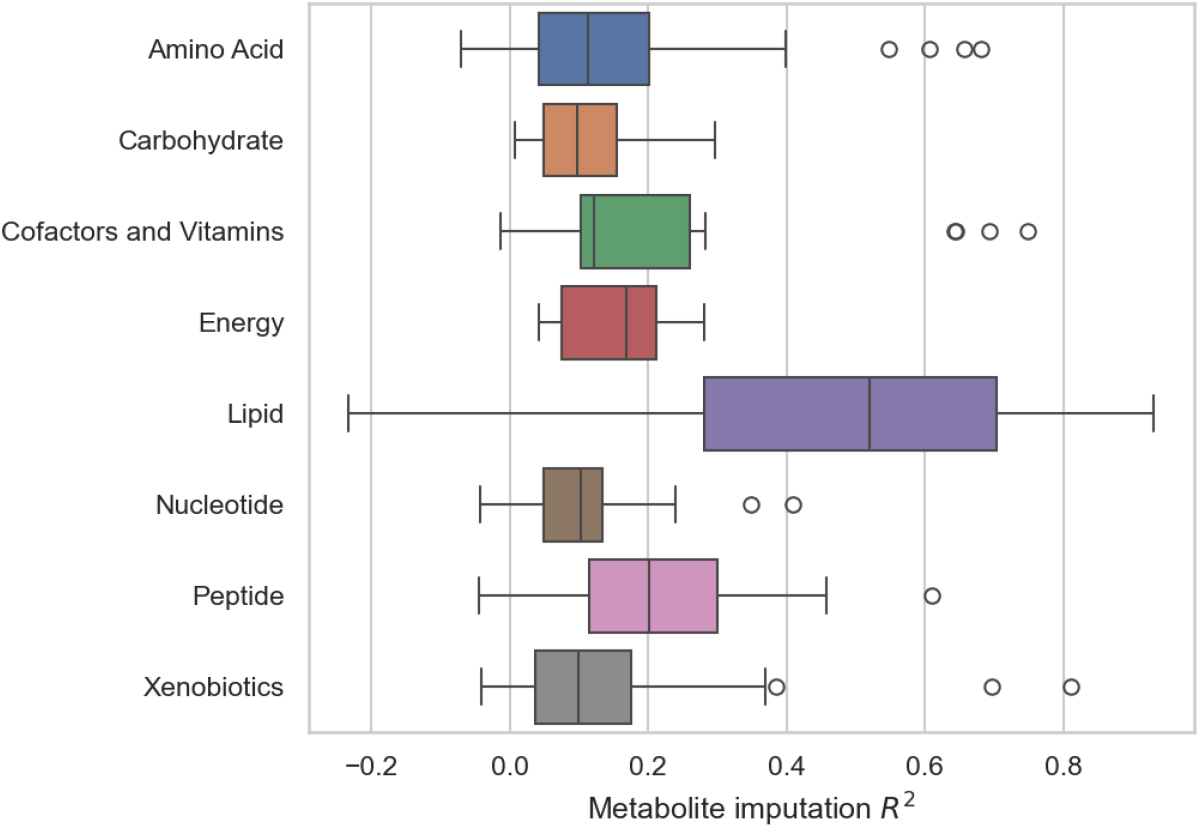
Imputation performance across metabolite compound classes based on Metabolon superclass classification.

After imputation of all Metabolon metabolites, the test set samples (n= 78) had a mean correlation of ρ = 0.61 (SD = 0.09, IQR 0.55-0.67), with the real Metabolon metabolites using the entire metabolic profile (**Figure 3c**). The metabolites that had an R^2^ ≥ 0.55, were taken forward for association studies (**Figure 3a**). Full details of the imputation model parameters and average performance on the training sets can be found in **Supplementary Table 2**. Individual Metabolon metabolite imputation performances on the test set can be found in **Supplementary Table 3**.

The study’s rigorous validation process involved comparing the imputed metabolites’ associations with BMI and CRP to those of actual Metabolon metabolites using data from the combined Metabolon-NPC and Metabolon-Only datasets (**Figure 5**). The beta coefficients were highly correlated between the real and imputed values with ρ = 0.93 and ρ = 0.89 for BMI and CRP respectively. Based on the standardised beta coefficient, the average association of real metabolites was 0.06 (SD = 0.12) with BMI and 0.01 (SD = 0.08) with CRP. The mean difference between the real and imputed metabolite associations was 0.005 (SD = 0.04) with BMI and 0.005 (SD = 0.04) with CRP. Out of the 199 metabolites tested, 10 metabolites (5% of all metabolites) had a beta coefficient difference greater than 1.96 standard deviations from the mean for BMI whilst CRP had 6 metabolites greater than 1.96 standard deviations (3% of all metabolites). The association comparison between real NPC LCMS metabolites with the imputed versions had similar concordances with a mean difference of -0.013 (SD = 0.04) for BMI and -0.019 (SD = 0.04) for CRP (**Figure 6**). For the four UK Biobank NMR metabolites which could be matched to the imputed Metabolon dataset (NPC-Only), the mean difference in association with the imputed metabolites was - 0.007 (SD = 0.05) and 0.01 (SD = 0.04) for BMI and CRP respectively. Full results of the association effect sizes for all models can be found in **Supplementary Table 4 and 5**.

**Figure 5.**
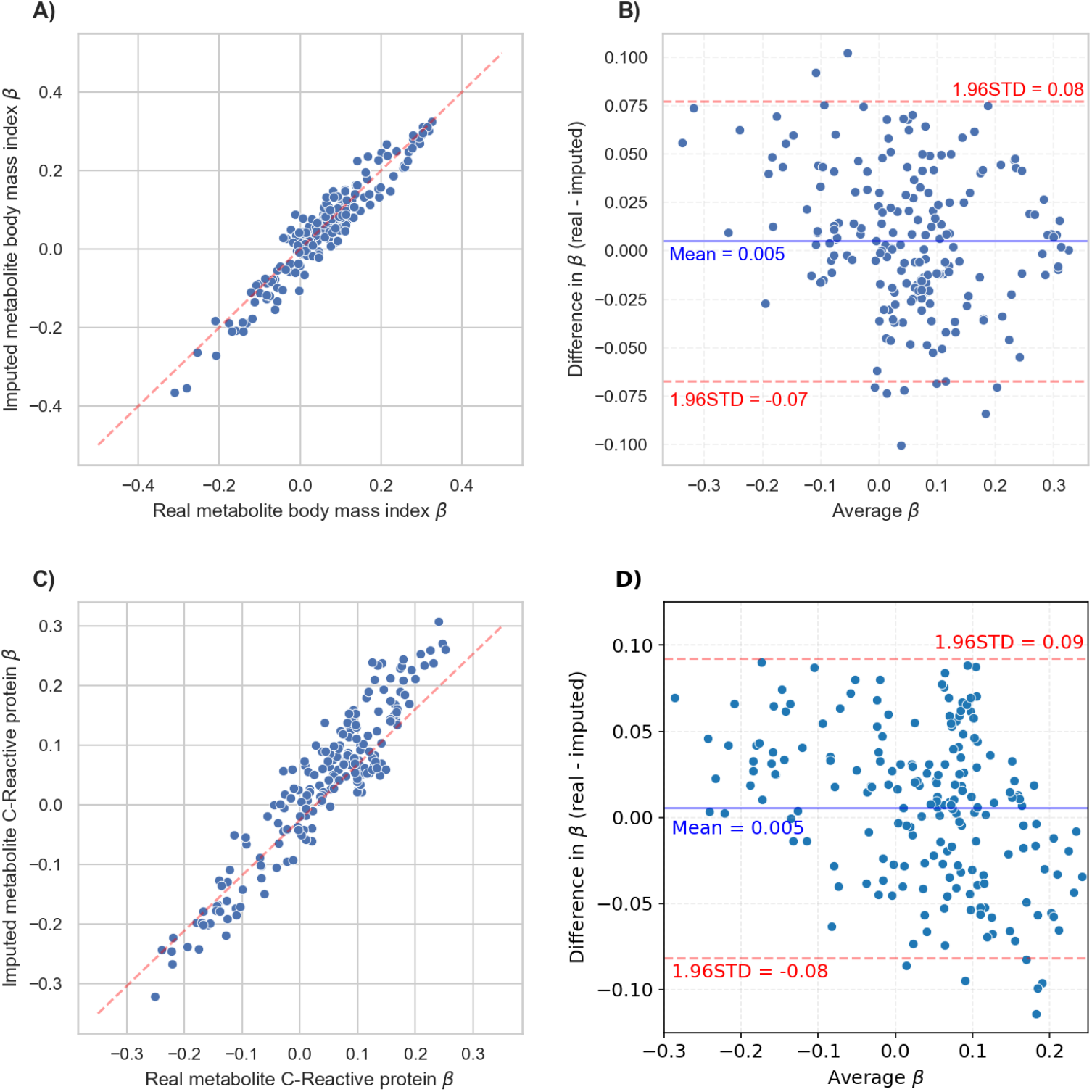
Comparison between metabolite associations with body mass index (BMI) a) real vs imputed effect size for BMI b) Bland-Altman plot of real and imputed effect size for BMI c) real vs imputed effect size for CRP d) Bland-Altman plot of real and imputed effect size for CRP

**Figure 6.**
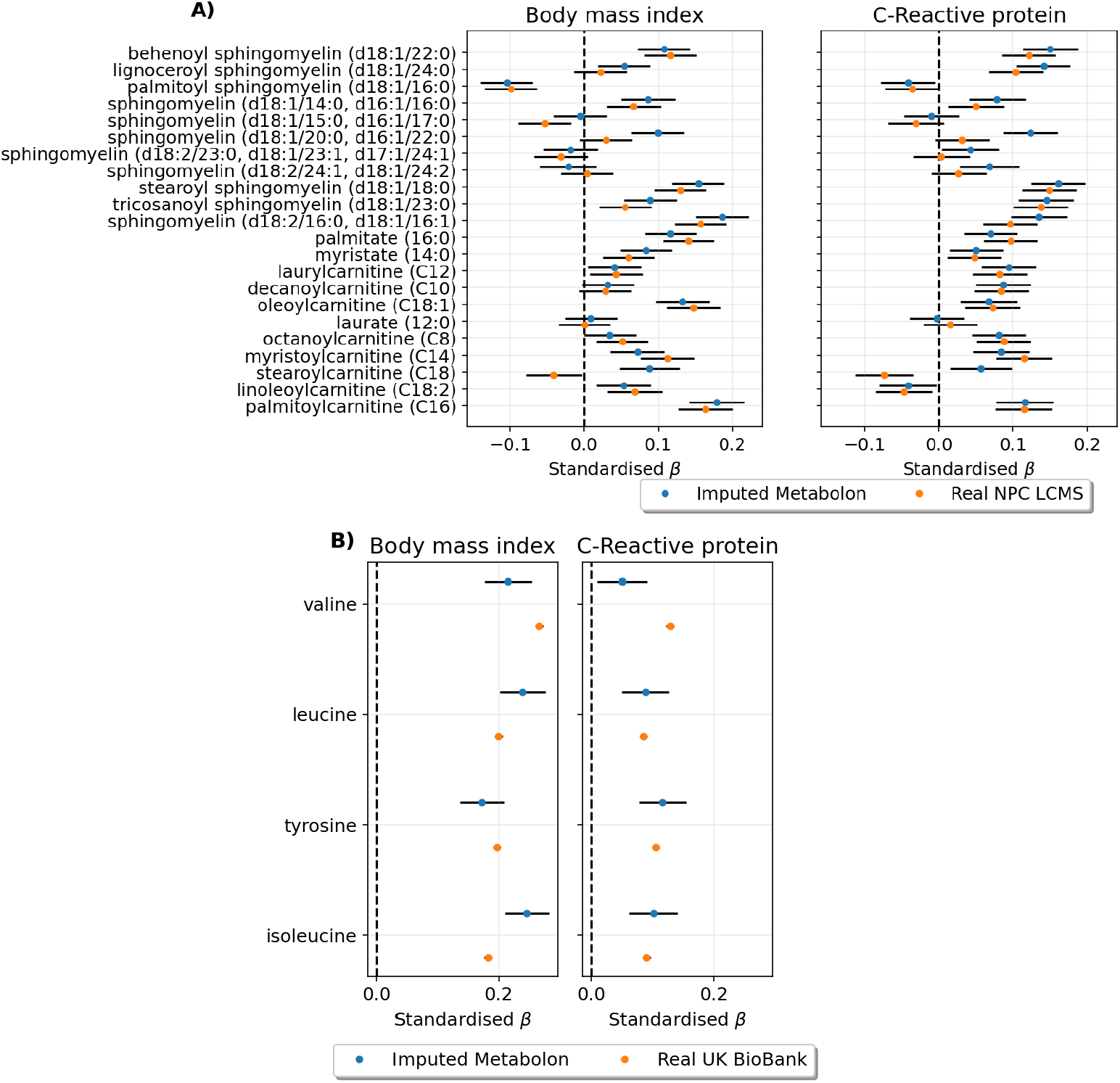
Comparison between metabolite associations with body mass index (BMI) and C-Reactive protein (CRP) using imputed Metabolon metabolites in the NPC-Only dataset and a) name-matched real NPC LC-MS b) real UK Biobank NMR

## Discussion

In this study, we applied deep learning techniques to train a model ensemble to generate an imputed Metabolon dataset comprising 199 metabolites, representing 22% of the metabolites in the original Metabolon dataset, using LC-MS metabolomic features from three platforms. We showed that the imputed dataset is expected to capture a minimum of 55% of the original variance (R^2^ ≥ 0.55) with low imputation uncertainty (R^2^ variance ≤ 0.025). Using our trained model, we generated an imputed Metabolon dataset of 199 metabolites for 2,971 samples which we combined with the Airwave Metabolon, Airwave NPC and UK Biobank datasets to run observational associations between metabolites and two important clinical outcomes: BMI and CRP. The results revealed strong alignment between associations observed using imputed Metabolon data and those found using real metabolites, underscoring the success of the model imputations.

The robustness of our approach was explored using a held-out test set to simulate the imputation of an external validation cohort. We evaluated various aspects of imputation performance, including overall accuracy (R^2^), model uncertainty (R^2^ variance), and correlation between imputed and real samples. The imputed dataset performed well across all metrics (Figure 3), confirming that our method is reliable for cross-platform imputation in external cohorts. To validate this further, we conducted a comparative observational analysis using the imputed dataset generated from an external cohort (NPC-Only) with three real metabolomics datasets. We selected two clinically relevant outcomes, BMI and CRP, which have well-documented metabolite associations^21^ to ensure a large set of significant metabolite associations where available for comparison. The concordance of associations for both outcomes across with all three real datasets and the imputed Metabolon dataset supported the reliability of our imputation approach (**Figure 5**).

The imputed Metabolon dataset primarily consisted of lipid metabolites (**Figure 4**), which was expected since the LC-MS platforms used for the imputation measure predominantly lipid compounds. In addition, given the relatively small sample size (N = 979), we constrained the size (number of layers and number of neurons) and training duration (early stopping) of our imputation models to mitigate the risk of overfitting. As a result, the imputation model trained on all metabolites and metabolomic features prioritized the identification of lipid imputing features over other metabolite classes. However, recognizing the importance of accurately imputing non-lipid metabolites, we expanded our imputation method to an ensemble which included models trained on each of the identified metabolite clusters, as depicted in **Figure 2**. The switch from a single model to an ensemble employing a greedy selection strategy significantly improved the imputation performance of non-lipid metabolites (**Figure 4)**. We anticipate that incorporating additional metabolomics platforms, such as NMR or reverse phase, would further enhance imputation across a broader range of metabolites, resulting in a more diverse imputation dataset.

The generative deep learning models implemented in this study have shown state of the art imputation for many data types such as imaging and RNA-seq data^22^. This success stems from the maximisation of a tighter lower bound of the observed data compared to other imputation methods, combined with their ability to make use of powerful multiple imputation methods with incomplete datasets. Our results demonstrate their extension to cross-platform metabolomics imputation by effectively identifying key features in an LC-MS dataset capable of inferring Metabolon metabolites. Our findings also highlight the potential application of IWAE for annotating metabolic features. By applying feature importance methods, such as SHapley Additive exPlanation (SHAP) values^23^, it would be possible to identify the specific LC-MS features used to infer each Metabolon metabolite. With this information, given that there seems to be a strong relationship between input features and imputed metabolites, the calculated feature importance could be used infer the annotation of unknown LC-MS features. However, further investigation into the feasibility and accuracy of this method for metabolomic feature annotation is warranted.

Our pioneering use of importance-weighted autoencoders to build an ensemble method for cross-platform imputation represents a meaningful step forward in untargeted LC-MS metabolomics for epidemiologic studies. Our method shows robustness and precision in generating imputed datasets for external cohorts which is crucial for downstream analyses, including associations with clinical outcomes. However, our study encountered limitations inherent to the LC-MS dataset used to impute Metabolon metabolites, resulting in suboptimal performance in the imputation of non-lipid metabolites. Additionally, the limited number of training samples necessitated restrictions on the deep learning model’s capacity and training time to prevent overfitting, which likely contributed to lower imputation performance.

Despite these limitations, our study highlights the potential of importance-weighted autoencoders and ensemble methods to improve cross-platform imputation and their potential application to assist with metabolomics feature annotation.

In conclusion, our study demonstrates the efficacy of using IWAEs to impute metabolomics data across different platforms, specifically between Metabolon and LC-MS datasets. The high concordance observed between real and imputed metabolites, particularly in associations with BMI and CRP, supports the robustness of this method. This approach offers a scalable solution to harmonize metabolomics data from diverse analytical platforms, facilitating data integration for replication studies and meta-analyses in large-scale epidemiological research.

## Data Availability

All data produced in the present study are available upon application and approval to the Airwave Health Monitoring Study.

